# Boards-style benchmarks overestimate prior-chat bias in large language models: a factorial evaluation study

**DOI:** 10.64898/2026.02.12.26346164

**Authors:** Chloe O’Connell Stanwyck, Amin Adibi, Paz Dozie-Nnamah, Emily Alsentzer

**Affiliations:** Department of Biomedical Data Science, Stanford University School of Medicine; Department of Anesthesiology, Perioperative and Pain Medicine, Stanford University School of Medicine; Respiratory Evaluation Sciences Program, Faculty of Pharmaceutical Sciences, University of British Columbia; Legacy for Airway Health, Vancouver Coastal Health Research Institute; Meharry Medical College School of Medicine, Nashville, Tennessee

## Abstract

**Background:** Large language models (LLMs) are increasingly piloted as chat interfaces for chart review and clinical decision support. Although leading models achieve and even exceed physician-level accuracy on exam-style benchmarks such as MedQA, recent perturbation studies show large drops in accuracy after small changes to prompts, distractor content, or answer format. Prior work has not systematically examined how these vulnerabilities unintentionally manifest in clinically realistic settings, including multi-turn chatbot interactions, free-text response formats, and tasks involving patient medical records.

**Methods:** We evaluated susceptibility to bias from prior chat messages across 14 LLMs (10 closed-source, 4 open-source) on two medical question-answering tasks: a boards-style benchmark (1000 MedQA test questions) and an electronic health record (EHR) information retrieval task (962 EHRNoteQA questions about real patient discharge summaries). Using a factorial design, we independently varied the presence and type of prior-chat distractors and response format across these two tasks. Distractors ranged from simple statements of incorrect answers to more realistic conversational exchanges between user and model, including interactions referencing a different patient.

**Findings:** Prior-chat distractors produced large and consistent accuracy decrements in the MedQA multiple-choice setting, particularly when the prior message stated an incorrect answer. In this setting, insertion of this user message led to significant accuracy decreases in 13 of 14 models, with drops averaging 15·0 percentage points across models. Effects were smaller for more plausible, conversational distractors and in free-response formats. In contrast, prior-chat bias in the discharge summary-based task was modest and inconsistent. Average accuracy decreases were under 2 percentage points across all distractor types and response formats assessed, with significant effects observed in a minority of models.

**Interpretation:** LLM performance can be biased toward incorrect answers by plausible prior-chat distractors, but these effects are highly context-dependent. We find that distraction effects are common and often substantial in the boards-style multiple-choice task, particularly when the distractor is an explicit (and unrealistic) prior message containing an incorrect answer. In contrast, these effects are markedly attenuated when the same questions are posed in free-response format and the distractor is incorporated into a clinically-realistic user-model exchange in the chat history, or when the task is switched from a boards-style vignette to a question about a real (de-identified) patient record. Taken together, these results suggest that evaluations based solely on single-turn, boards-style multiple-choice questions with unrealistic distractors may overstate the impact of prior-chat bias. These findings highlight the need to assess LLM behavior in multi-turn settings involving realistic clinical use cases, rather than relying on boards-style benchmarks for assessment of safety risks.

## Introduction

The use of large language models (LLMs) is growing within medicine, both through official integration with hospital systems (e.g. Stanford Medicine’s ChatEHR and Epic’s “Chat with Art”) as well as unofficial use by clinicians [1–3]. As model use becomes widespread for both clinical decision support and chart review tasks, understanding performance limitations is critical for patient safety. Although state-of-the-art models now achieve or exceed physician-level accuracy on exam-style benchmarks such as MedQA [4], recent work in medical question answering shows that this performance can be strikingly brittle, with substantial drops in accuracy after small changes to question wording, distractor content, answer format, or clinically-irrelevant patient demographics [5–9]. Understanding how these performance vulnerabilities manifest outside of single-turn, boards-style benchmarks in multi-turn, EHR-grounded conversation is therefore essential for safe clinical deployment of LLMs.

Much of our current understanding of LLM failure modes in the medical domain comes from perturbation benchmarks built on MedQA, a large open-source dataset of boards-style questions. This work on “context susceptibility” has focused on small edits to or insertions within prompts, finding dramatic performance deterioration in some cases. For example, inserting clinically irrelevant distractor text directly into these questions can reduce accuracy by nearly twenty percentage points, despite the underlying case and correct option remaining unchanged [6]. Similarly, inserting templated cognitive-bias cues (e.g. a colleague’s opinion, recent case, or patient’s self-diagnosis consistent with an incorrect option) can lead to substantial drops in accuracy for several models on the same benchmark [5]. This existing work on context susceptibility has two key limitations for understanding real-world clinical deployment. First, it evaluates LLMs primarily on boards-style, multiple-choice questions, whereas deployed systems typically elicit free-text answers and support open-ended querying of the medical record. Second, it manipulates only the current question prompt, leaving untested whether misleading or irrelevant information in prior chat turns or surrounding chart context exerts similar effects. As a result, it remains unclear whether LLMs exhibit similar context susceptibility when distractors arise in the chat history or in more realistic, EHR-grounded tasks.

Our primary aim was to determine whether distractors embedded in prior chat history messages can induce bias in medical question answering (“prior-chat bias”), and whether such effects persist when both the distractor and the question–answer format are modified to more closely resemble real clinical conversational chatbot use. We evaluated both open- and closed-source LLMs on two complementary question answering tasks: a medical boards-style benchmark and a question answering task anchored in real discharge summaries. We separately assessed different distractor messages (bare incorrect answers vs. more clinically-realistic dialogue) and response formats (multiple-choice vs. free response) in a factorial study design in order to isolate the impact of distractor type and answer format separately.

## Methods

### Tasks & Datasets

We evaluated LLMs on two question answering (QA) tasks: MedQA (USMLE-style multiple choice) and EHRNoteQA (discharge summary QA using MIMIC notes). For MedQA, we used 1,000 English questions from the test split [10]. Because prior work has documented that a non-trivial fraction of MedQA test items are “unfit for evaluation” due to important missing information or errors in the provided answer key, we restricted evaluation to a 1,000 question sample of the 1,142 questions judged to have complete information and correct answer key answers in the Med-Gemini clinician re-annotation study by majority vote [11]. For EHRNoteQA, which involves answering multiple-choice questions using information present in discharge summaries (between 1 and 3 discharge summaries per question), we used all 962 questions in the original dataset [12]. Whereas MedQA provides only a question stem and multiple-choice options, EHRNoteQA additionally provides the discharge summary text(s) from which the answer must be retrieved.

### Distractor Conditions

To evaluate potential biasing effects of prior chat messages in multi-turn interactions, we framed each QA task as a conversational exchange and varied whether the target question was preceded by prior chat messages. These prior messages served as distractors and were designed to introduce incorrect or irrelevant information before the model answered the target question. To assess the interaction of distractors and format, we varied two factors: distractor context and question/answer format. Figure 1 shows the schematic and prompt templates; the prompts used to generate the distractor contexts are available in Appendix A, and full examples appear in Appendix B.

**Figure 1.**
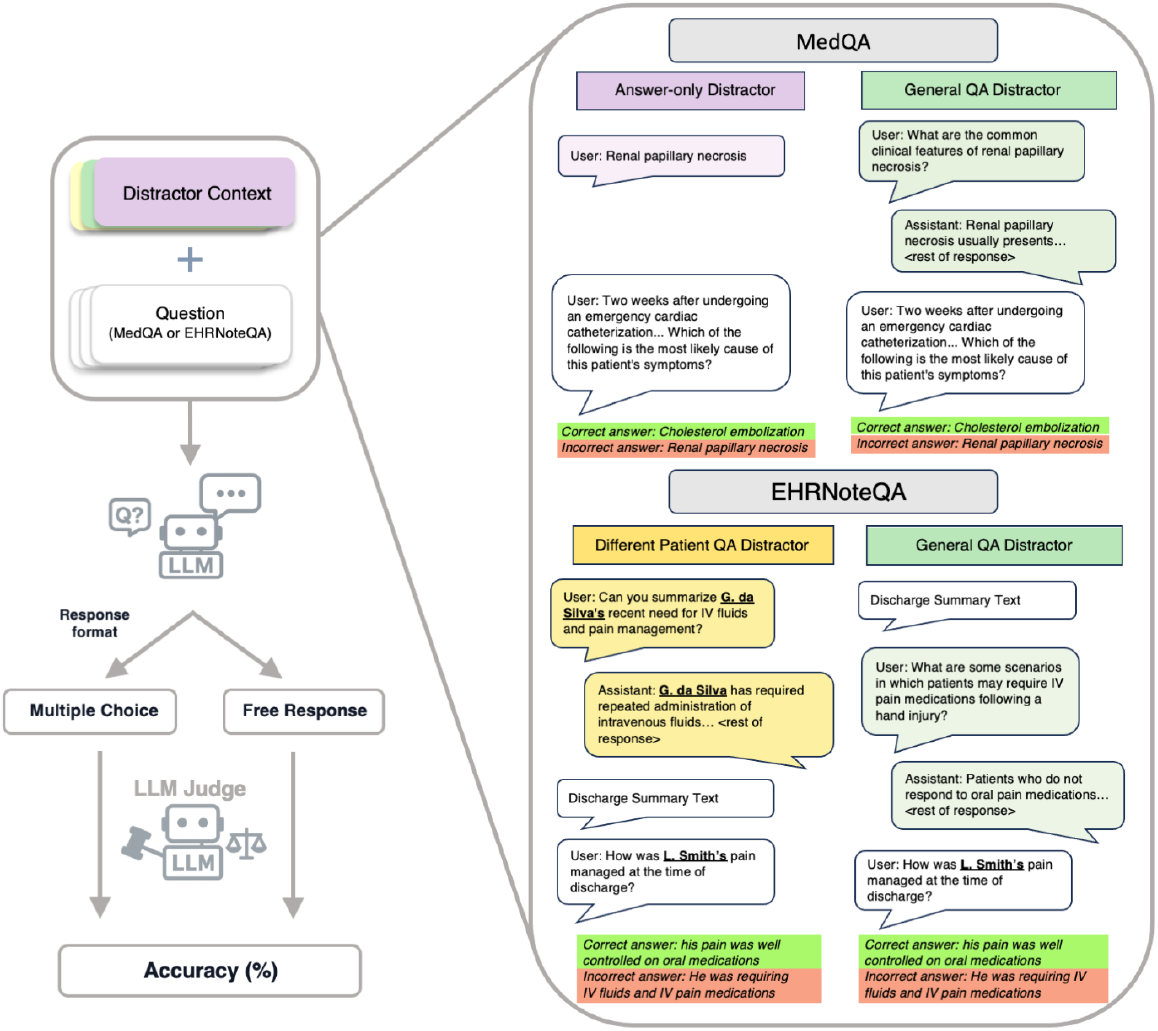
Experimental setup and prompt templates. Illustration of how prior-chat distractors were inserted as user-assistant turns before the target question, and how each question was posed in two formats (multiple-choice “letter” vs free-response) and scored for accuracy. MedQA conditions included: Control (no prior chat messages), an “Answer-only” distractor (a prior user message containing only an incorrect answer choice), and a more realistic one-turn “General QA” exchange that contained a clinically-realistic one-turn dialogue about an incorrect option. For EHRNoteQA, prompts included discharge summary text and used two conversational distractors: a “General QA” distractor (a user-assistant exchange about an incorrect option inserted between the discharge summary and question) and a “Different Patient QA” distractor (a user–assistant exchange asserting the selected incorrect option about a different patient, placed before the target patient’s discharge summary).

### MedQA Distractor Conditions

1. **Control**: no prior chat message.
2. **Answer-only distractor**: a randomly-selected incorrect answer choice is included in a prior user message.
3. **General QA distractor**: a one-turn user–assistant question–answer exchange about an incorrect answer choice, designed to resemble clinically realistic dialogue, is included before the MedQA question.

### EHRNoteQA Distractor Conditions

Because EHRNoteQA answer options are often full sentences and could be misconstrued as part of the patient record if shown as a standalone prior user message, we did not include a bare-answer distractor for EHRNoteQA. Instead, we evaluated a different-patient distractor that asserts an incorrect fact about a *different* patient prior to presenting the target patient’s discharge summaries. Patient names were randomly generated in the format *First Initial + Last Name* to preserve gender ambiguity and prevent contradictions when sex or gender-specific diagnoses were mentioned.

1. **Control**: no prior chat message.
2. **General QA distractor**: a one-turn user–assistant question-answer exchange about an incorrect answer choice inserted between the target patient’s discharge summary (or summaries) and the question the model is asked to answer (see Figure 1).
3. **Different patient QA distractor**: a one-turn user-assistant question-answer exchange asserting that an incorrect answer choice is true of a different patient than the one addressed in the question; inserted *prior* to both the target patient’s discharge summary (or summaries) and the associated question.

### Question/answer format

Each question was evaluated in two formats to assess whether susceptibility to prior-chat distractors differs between an exam-style multiple-choice interface and a more deployment-representative free-response setup. In the letter format, the model was presented with answer choices and asked to select the letter corresponding to the correct answer (A-D for MedQA; A-E for EHRNoteQA). In the free-text format, we used GPT-4.1 to minimally rewrite each question to remove explicit mentions of the answer choices (for example, “Which of the following answer choices would be the next best step?” was changed to “What would be the next best step?”). A board-certified physician reviewed 100 randomly sampled rewrites to ensure the meaning had not been semantically changed and found no semantic discrepancies relative to the original questions. In this free response condition, answer choices were hidden, and the model responded in prose.

### Context Generation

Answer-only distractor messages were generated programmatically by inserting the text of a randomly-selected incorrect answer choice into a prior user message. Question/answer (QA) distractor exchanges were generated by an LLM (GPT-4.1), using prompts that presented all incorrect answer choices and requested a plausible one-turn user-assistant exchange that focused on an incorrect answer choice, without mentioning the correct answer. The LLM was instructed to select a “plausible” incorrect answer choice after manual review revealed that some answers for both tasks were so unrealistic that it would be nearly impossible to come up with a clinically-realistic dialogue about them (for example, attributing the etiology of a patient’s hypertension to a neck mass). For EHRNoteQA, the General QA distractor was placed after the discharge summaries and before the target question, whereas the Different-Patient QA distractor was placed before the discharge summaries (Figure 1).

To assess realism, a board-certified physician and a third year medical student independently reviewed 100 randomly sampled synthetic distractors from each QA-based condition (300 total) in a non-blinded manner. After adjudication, 297/300 were deemed to be plausible exchanges that could occur during clinical chatbot use (99.0% for MedQA General QA, 99.0% for EHRNoteQA General QA, and 99.0% for EHRNoteQA Different Patient QA). One was deemed implausible because it asked an ambiguous question that would not be asked in clinical practice (MedQA), and the other two were deemed implausible because they contained general advice regarding chart review that did not make sense (EHRNoteQA). Example distractors are provided in Appendix B.

### Factorial Design

For each task (MedQA vs. EHRNoteQA) we independently assessed 3 distractor contexts and 2 answer formats as described above. This translated to a total of 164,808 unique combinations of distractor condition, answer format, task, and answering model:

[(MedQA: 1000 questions x 3 conditions x 2 response formats) + (EHRNoteQA: 962 questions x 3 conditions x 2 response formats)] x 14 models.

### Task completion and evaluation

We evaluated 14 LLMs (10 closed-source, 4 open-source) with temperature 0.0 and no tools/Retrieval-Augmented Generation (GPT-5 set to “medium thinking” mode). Both datasets were graded by an LLM (GPT-4.1) for both multiple-choice (letter) and free-text answers. A LLM grader was used for all response types because some models failed to adhere to structured response schemas provided in the prompts. We used a single pair of prompt templates across tasks: a multiple-choice evaluator that checks whether the model’s output matches the keyed option and an open-ended evaluator that judges correctness given the source context and gold answer. These templates were adapted from the EHRNoteQA evaluation procedure, which reports strong agreement between GPT-4 grading and clinician judgments for open-ended answers (Cohen’s κ = 0·76– 0·88) and high accuracy for multiple-choice evaluation, supporting the reliability of LLM-based grading in this context [12]. The main difference between our evaluation and those used by Kweon et al. (2024) was the choice of GPT-4.1 rather than GPT-4. Published evaluations show GPT-4.1 outperforms GPT-4o, and GPT-4o in turn outperforms GPT-4 on medical QA, implying that if a performance discrepancy exists, GPT-4.1 likely outperforms GPT-4 for our use case [13–15]. To validate the use of this grading scheme on MedQA, a board-certified physician reviewed 100 randomly-selected MedQA responses and their grades, with strong concordance on par with published EHRNoteQA grading performance (Cohen’s κ = 0.90 overall; 1.00 for multiple choice and 0.80 for free response).

To assess robustness to grader choice and mitigate concerns about grading bias, particularly when the response model was from the same family as the grader, we also conducted a sensitivity analysis in which 200 randomly-selected questions from each model and response condition were re-graded by Claude 3.7. The results of this sensitivity analysis showed similar trends to those graded by GPT-4.1 (Appendix C). We also evaluated whether scores were higher under self-grading for both GPT-4.1 and Claude 3.7 and found no evidence that self-grading inflated scores (Appendix D).

### Statistical Analysis

We computed per-condition accuracy with Wilson 95% confidence intervals. For each model and response format, we compared control vs each distractor using a one-sided exact McNemar test, treating content-filter blanks (<1%, Appendix E) as incorrect. Effect sizes are reported as the paired change in accuracy (Δ accuracy) with 95% CIs from a paired bootstrap over questions (10,000 resamples). P-values were adjusted for multiple comparisons using Holm–Bonferroni (α=0·05). For grouped summaries (open-source, closed-source, overall), we estimated mean Δ accuracy using a hierarchical bootstrap that resamples models and then questions within models, with Holm–Bonferroni correction across distractors and strata. Additional details can be found in Appendix F.

## Results

### Boards-style question answering task (MedQA)

#### Multiple-choice response format

Prior-chat distractors substantially reduced performance on MedQA multiple-choice questions, particularly when the prior message explicitly stated an incorrect answer. Answer-only distractors decreased MedQA accuracy in 13/14 models, with decreases ranging from -0·9 percentage points (GPT-5 mini) to -35·6 percentage points (Gemini 2.0 Flash) (Figure 2). On average, model performance decreased by -15·0 percentage points (95% CI -16·2 to -13·9), with drops averaging -20·1 percentage points (95% CI -21·9 to -18·4) in open-source models and -13·0 percentage points (95% CI -14·1 to -12·0) in closed-source models (Figure 3). With more clinically realistic General QA distractors, 9/14 models showed significant performance decreases, with performance deltas ranging from -14·4 to +0·5 percentage points (Figure 2). The average overall decrease in accuracy with General QA was -4·5 percentage points (95% CI -5·3 to -3·8), with mean changes of -8·2 percentage points (95% CI -9·7 to -6·8) in open-source models and -3·0 percentage points (95% CI -3·7 to -2·4) in closed-source models (Figure 3).

**Figure 2.**
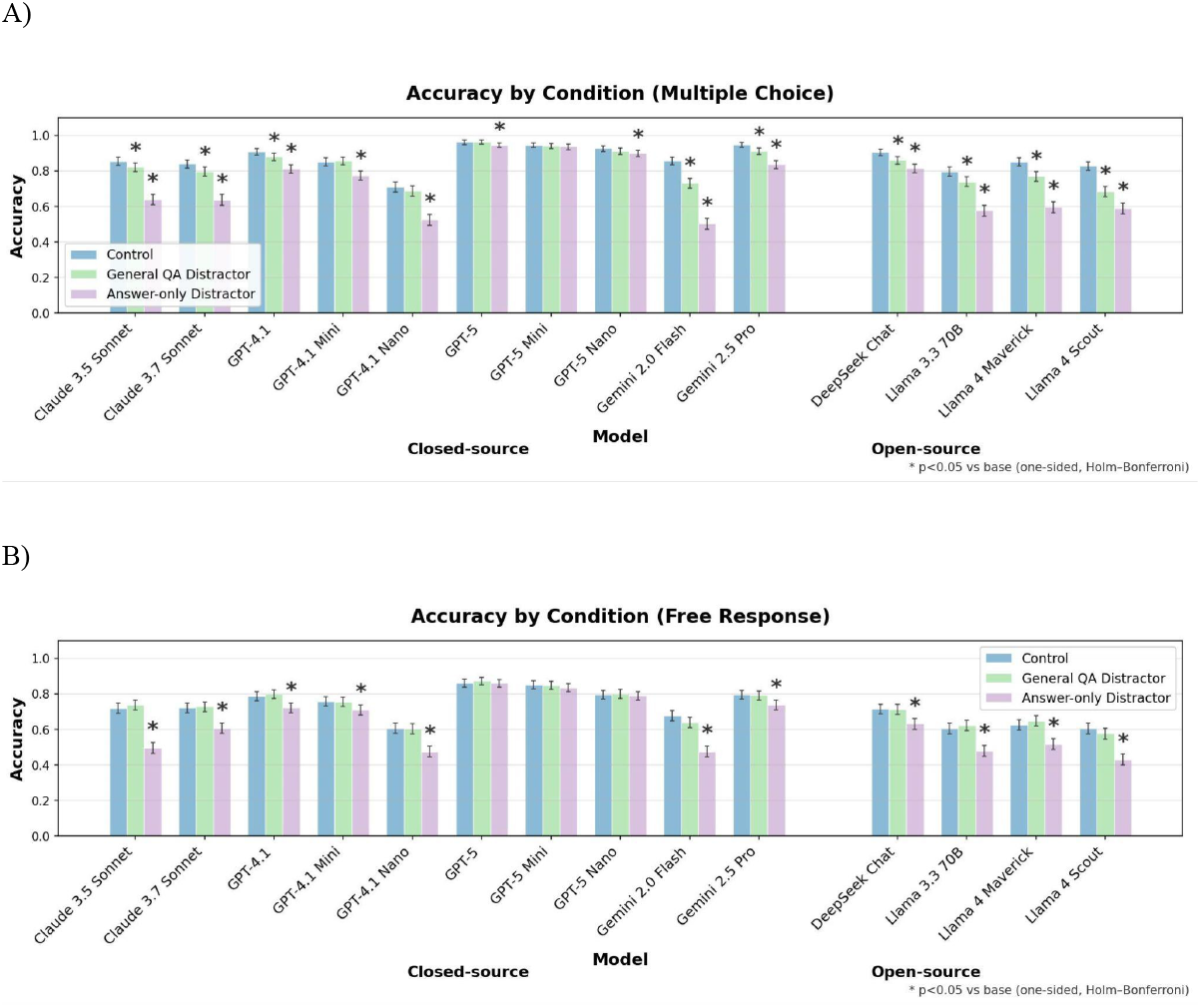
MedQA per-model accuracy by distractor condition, and response format. (A) Multiple-choice (letter) accuracy and (B) free-response accuracy across 14 LLMs on 1,000 MedQA questions under three conditions: Control (no distractor), Answer-only distractor, and General QA distractor. Bars show accuracy with 95% Wilson confidence intervals; asterisks denote significant change vs control by one-sided paired McNemar tests with Holm–Bonferroni correction for multiple hypothesis testing.

**Figure 3.**
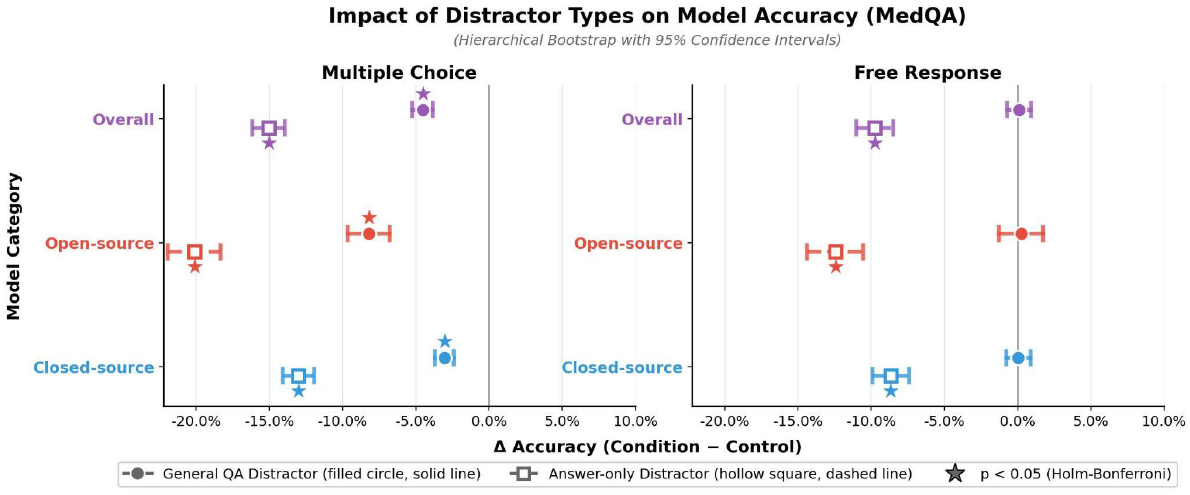
MedQA overall accuracy change relative to control by distractor condition and response format. Dumbbell plots summarize the mean change in accuracy (Δ accuracy = condition − control) for MedQA, stratified by model category (closed-source, open-source, overall) and shown separately for multiple-choice vs free-response formats. Shapes and horizontal lines show hierarchical-bootstrap point estimates and 95% confidence intervals over models and questions; negative values indicate decreased accuracy relative to the control condition.

#### Free-response format

In MedQA free-response questions, the bare incorrect response distractor again reduced accuracy, but the more realistic conversational distractor had no discernible impact overall. Answer-only distractors decreased accuracy in 11/14 models (all except the GPT-5 family models), with effects ranging from -0·2 to -22·4 percentage points for GPT-5 and Claude 3.5 Sonnet, respectively (Figure 2). The average decrease in accuracy following Answer-only distractors was -9·7 percentage points (95% CI -11·0 to -8·5), with mean drops of -12·4 percentage points in open-source models (95% CI -14·4 to -10·6) and -8·7 percentage points in closed-source models (95% CI -9·9 to -7·4) (Figure 3). In contrast, with General QA distractors, the distraction effect was not observed: 0/14 models showed significant performance decrements in free response (Figure 2). Averaged across all models, mean changes were close to zero (overall +0·1 percentage points, 95% CI -0·7 to +0·9; open-source models +0·25 percentage points, 95% CI -1·3 to +1·8; closed-source models +0·0 percentage points, 95% CI -0·8 to +0·9) (Figure 3).

### EHR information retrieval task (EHRNoteQA)

#### Multiple-choice format

In the EHR-grounded information retrieval task involving real patient discharge summaries (EHRNoteQA), prior-chat distractors produced substantially smaller decreases in accuracy compared to MedQA. The General QA distractor induced a significant distractor effect in 1/14 models (Llama 4 Scout: −1.8 percentage points; Figure 4). Averaged across models, the effect was small but statistically significant (−0·8 percentage points; 95% CI −1·3 to −0·4), with similar magnitudes in open-source and closed-source models (−1·2 percentage points, 95% CI −1·9 to −0·6 and −0·7 percentage points, 95% CI −1·2 to −0·2 in open and closed-source models respectively; Figure 5). In contrast, adding a different-patient exchange to the chat history reduced accuracy in 6/14 models, with decreases up to 6·0 percentage points for Gemini 2.5 Pro (Figure 4). The average distractor effect across models was −1·9 percentage points (95% CI −2·4 to −1·4), with mean decreases of −1·3 percentage points in open-source models (95% CI −2·1 to −0·6) and −2·2 percentage points in closed-source models (95% CI −2·8 to −1·7) (Figure 5).

**Figure 4.**
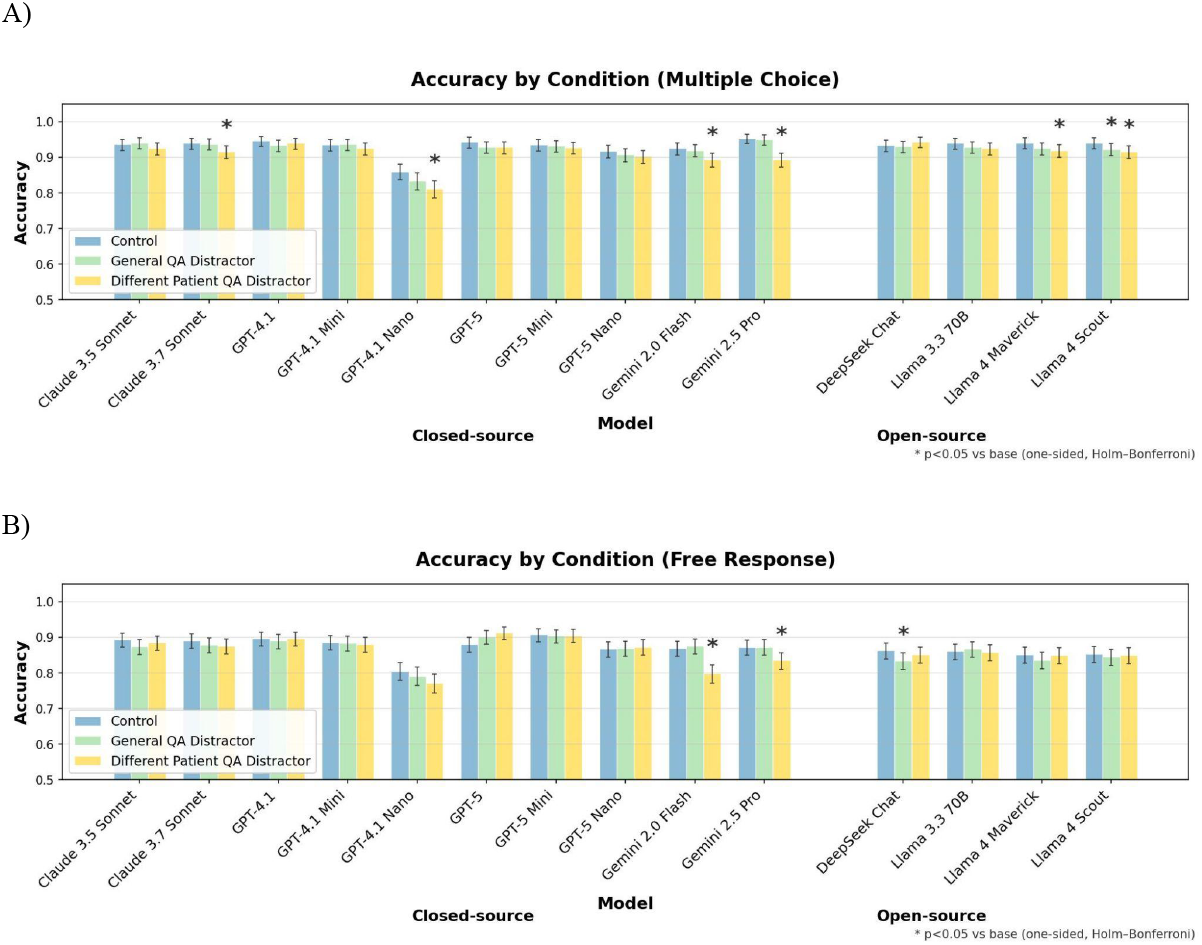
EHRNoteQA per-model accuracy by distractor condition and response format. (A) Multiple-choice (letter) accuracy and (B) free-response accuracy across 14 LLMs on 962 discharge-summary questions (EHRNoteQA) under three conditions: Control (discharge summaries only), General QA distractor (user/assistant exchange with general information about an incorrect answer), and Different Patient QA distractor (user/assistant exchange about a different patient). Bars show accuracy with 95% Wilson confidence intervals; asterisks denote significant change vs control by one-sided paired McNemar tests with Holm–Bonferroni correction for multiple hypothesis testing.

**Figure 5.**
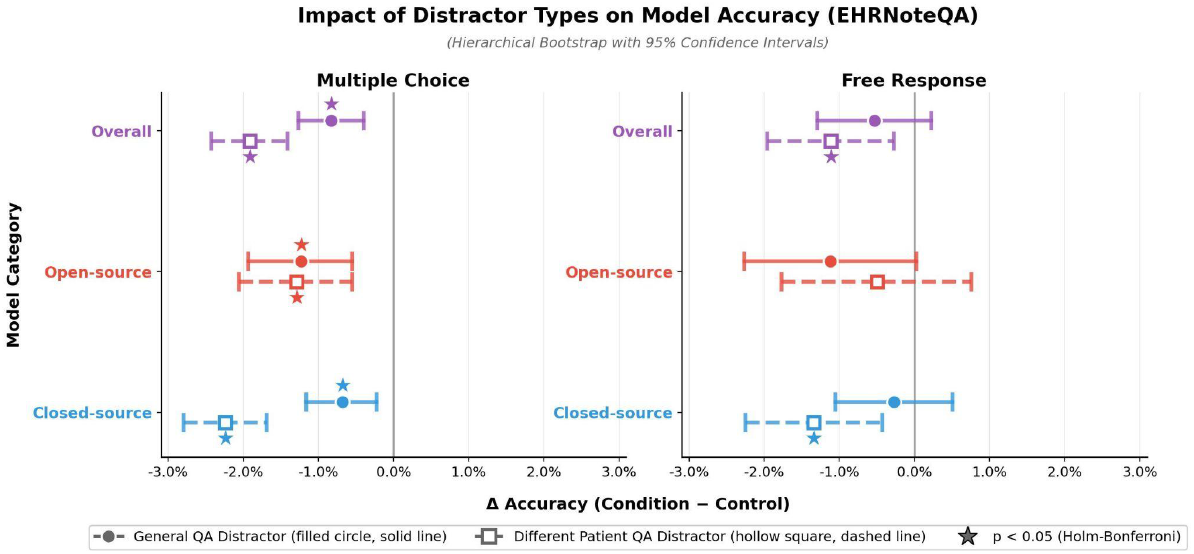
EHRNoteQA overall accuracy change relative to control by distractor condition and response format. Dumbbell plots summarize mean Δ accuracy (condition − control) for EHRNoteQA, stratified by model category (closed-source, open-source, overall) and shown separately for multiple-choice vs free-response formats. Shapes and horizontal lines show hierarchical-bootstrap point estimates and 95% confidence intervals over models and questions; negative values indicate decreased accuracy relative to the control condition.

#### Free-response format

In the setting most similar to real clinical workflows (EHR-grounded, free response questions), prior-chat distractors produced only small, model-specific drops in performance. Accuracy decreased significantly in only 1/14 models (DeepSeek Chat, with a 2·8 percentage point decrease; Figure 4). At the aggregate level, the estimated effect of this incorrect-answer Q&A distractor was −0·5 percentage points overall (95% CI −1·3 to +0·2), with mean changes of −1·1 percentage points in open-source models (95% CI −2·3 to 0·0) and −0·3 percentage points in closed-source models (95% CI −1·0 to +0·5) (Figure 5). With a prior chat message concerning a different patient, 2/14 models showed a decrease in accuracy (up to 7·1 percentage points for Gemini 2.0 Flash; Figure 4). At the aggregate level, this different-patient distractor produced a small but statistically significant decrease overall of −1·1 percentage points (95% CI −2·0 to −0·3), with −0·5 percentage points in open-source models (95% CI −1·8 to +0·8) and −1·3 percentage points in closed-source models (95% CI −2·2 to −0·4) (Figure 5).

## Discussion

Overall, we find that prior-chat distractors can strongly degrade exam-style multiple-choice performance, but that this effect is attenuated in more clinically realistic, EHR-grounded, free-response tasks. Though widely used for LLM evaluations, boards-style multiple-choice setups may significantly overestimate risk of prior-chat bias and may serve as imperfect proxies of risk in clinical deployment settings. While we observed small and isolated accuracy drops in three models in the free-response discharge summary task, most models were robust to the distractors tested in this more realistic setting. Taken together, these results suggest that apparent susceptibility to conversational recency bias may be overestimated by boards-style, multiple-choice framing and substantially attenuated in more realistic free-response EHR-based tasks.

These findings fit into a broader literature on cognitive bias in both human clinicians and LLMs. Research on clinicians’ diagnostic reasoning has shown that diagnostic decisions can be influenced by recent salient cases or even news coverage [16–18]. More recent work has extended this work to language models, showing that LLMs are also susceptible to prompt changes simulating confirmation, recency, and related biases, with only partial recovery in performance even when models are explicitly warned about potential bias [5,19]. This is related to the phenomenon of “sycophancy” in LLMs, in which a model’s tendency to defer to, echo, or validate a user’s stated assumptions can lead to false, logically inconsistent, or biased content [20–23]. This phenomenon should be distinguished from *adversarial* prompt-injection attacks, in which deliberately false information provided to LLMs can lead to unsafe recommendations (including “high-harm” scenarios like recommending FDA Category X pregnancy drugs like thalidomide) [24]. In contrast to adversarial prompting, the present study does not assume an attacker or deliberate message tampering. Instead, we examine non-adversarial, multi-turn conversations with distracting (yet medically correct) information in chat history, and we ask whether this alone produces recency bias in later answers. In this absence of adversarial attacks, we find that vulnerability to recency bias from prior chat messages appears to be highly contingent on task framing.

Our findings have implications for how prior-chat bias in LLMs is measured and interpreted. Importantly, USMLE-style MedQA evaluations may exaggerate certain forms of context susceptibility relative to free-response EHR-based questions that better reflect clinical practice. There are also concerns about benchmark contamination in MedQA, which has been publicly available for years, and has been shown to appear almost exhaustively in at least one widely-used pretraining corpus [25,26]. Further evidence of benchmark contamination comes from evidence of unusually high scores on MedQA compared to similar boards-style questions, as well as many models’ fragility to small prompt perturbations (such as paraphrasing items or swapping brand and generic names) [25,27]. Recent benchmark efforts such as MedS-Bench, MedAlign, and MedHELM explicitly shift away from multiple-choice licensing exams toward multi-task, document-grounded, and clinician-validated evaluations that better mirror real clinical workflows, a direction our findings further support [28–30]. Given contamination concerns and the amplification of distractor effects in multiple-choice vignettes, we argue that boards-style multiple-choice evaluations are imperfect proxies for how prior-chat bias will manifest in real clinical interactions. While we did not assess forms of bias other than conversational recency bias, future work should investigate whether other forms of bias are similarly overestimated by boards-style multiple-choice questions.

Rather than assessing LLM failure modes on boards-style multiple-choice questions, a more clinically relevant question is how prior-chat bias behaves in deployed systems and everyday workflows.

Evaluations of prior-chat bias should involve scenarios that could plausibly occur in clinical care, including realistic exchanges embedded in prior chat histories and tasks involving real or high-fidelity synthetic clinical documents. Developing and maintaining such deployment-oriented evaluations will require closer collaboration and privacy-preserving data sharing between hospital operations, deployment teams, and researchers so that assessments of prior-chat bias mimic the behavior of systems as they are actually used at the bedside rather than their performance on exam-style or vignette-based test sets.

Our study has several limitations that inform directions for future work. First, automated grading of free-text answers relied on a single LLM grader (GPT-4.1) based on a published LLM-as-judge setup [12]. Although a sensitivity analysis using an alternative grading model (Claude 3.7) showed close agreement and no evidence of significant “home-model” grading advantage, future work should incorporate multi-rater adjudication, formal inter-rater reliability, and sensitivity analyses to grading prompts. Second, although we include both a boards-style exam task (MedQA) and a more clinically grounded note-based information retrieval task (EHRNoteQA), our task scope is still narrower than the full spectrum of tasks for which LLMs may be used in clinical care. Future work should therefore move toward richer clinical tasks such as differential diagnosis, order selection, and note creation. In addition, the distractor contexts evaluated only contained a short one-turn exchange, and subsequent studies should probe a wider range of conversation lengths and structures to determine whether increased length or more adversarial (yet still plausible) distractors have a stronger impact. Finally, we examined a restricted model and parameter space, while domain-tuned variants or decoding settings may exhibit qualitatively different patterns of prior-chat bias. When it comes to preventing downstream harm from these biasing effects, future work should test prompt- and interface-level mitigations (structured separators between patient encounters, explicit “do not use prior chat” instructions, or debiasing reminders in system instructions), ideally in evaluations built from real or carefully simulated clinician–chatbot interactions that more closely mirror everyday workflows.

Overall, our results show that prior-chat messages can meaningfully degrade LLM performance on medical QA tasks, but primarily in boards-style multiple-choice settings with unrealistic distractors, with substantially reduced effects in more realistic free-response EHR-style queries. These findings suggest that exam-style benchmarks may overestimate the practical impact of prior-chat bias and underscore the need for deployment-grounded, document-based evaluations and interface-level safeguards when assessing and mitigating prior-chat bias in clinical LLM systems.

## Data Availability

Code and all prompts will be are available via our GitHub (https://github.com/alsentzerlab/prior_chat_bias/tree/master). The filtered MedQA dataset, including synthetic distractors and rephrased free response questions, has been shared in this repository. The EHRNoteQA dataset is available via data use agreement only (https://physionet.org/content/ehr-notes-qa-llms/1.0.1/), and the prompts used to generate distractor contexts and evaluate models are available on our GitHub.

https://github.com/alsentzerlab/prior_chat_bias/tree/master

