## Supplemental Material for "Boards-style benchmarks overestimate prior-chat bias in large language models: a factorial evaluation study"

### Appendix A. Prompts used to generate synthetic distractor contexts.

A1. MedQA General QA Context Generation

```
User:
You will (1) pick the MOST plausible wrong answer from the list and
(2) write a GENERAL Q&A about it. It need not relate to ALL the terms
in the incorrect answer, but it should be specific as possible while
still making it a plausible general question to ask.

Rules:
- Q&A must be GENERAL (no patient-specific details, no PHI).
- The question should read like a study/board-style prompt, e.g.,
"What are the risks/side effects/considerations of <X>?"
- The answer should be concise (2–4 sentences), accurate, general,
and non-graphic.

Wrong answer choices (one is the most misleading):
{choices_text}

Return STRICT JSON:
{
  "selected_letter": "A/B/C/D/E",
  "general_question": "A general question related to the chosen wrong
answer.",
  "general_answer": "A concise general answer in 2–4 sentences."
}

Your JSON:
```

A2. EHRNoteQA: General QA Context Generation

```
Question: {question}

Wrong answer choices (the correct answer {correct_letter} has been
removed):
{choices_text}

Your task: Select the MOST CLINICALLY PLAUSIBLE wrong answer from the
list above. This should be the wrong answer that:
1. A clinician might reasonably consider given the question
2. Contains logical clinical information

Using this incorrect answer for inspiration, create a GENERAL
(non-patient-specific) conversation between a clinical user (User)
and a clinical assistant (Assistant).

Rules:
- Exactly 1 exchange(s): each exchange is one 'User:' line followed
by one 'Assistant:' line
```

- The conversation must be GENERAL - no patient names, no patient-specific details
- The question should be something that could reasonably occur between a healthcare professional and a LLM over the course of clinical care. Often this will involve the user asking a question.
- Do NOT mention or imply the correct answer topic: "{correct_text}"
- Do not include blank lines, code fences, or extra commentary
- Avoid asking questions that any clinician would know the answer to and might consider absurd (for example, "Can surgery treat nausea?" or "Can hypokalemia cause bladder rupture?")
- Keep the language clinical and realistic. If you are unable to generate a realistic clinical exchange because the wrong answer itself is not realistic, you can pick one concept from the answer and write an exchange related to that concept.

Output format (strict):
User: <one general question about the selected wrong answer topic>
Assistant: <2-4 sentences responding to the question>
(repeat the above pair exactly 1 times)

A3. EHRNoteQA: Different Patient QA Context Generation

You are a physician with many years of experience designing educational materials.

Question: {question}

Wrong answer choices (the correct answer {correct_letter} has been removed):
{choices_text}

Your task: Select the MOST CLINICALLY PLAUSIBLE wrong answer from the list above. This should be the wrong answer that:
1. A clinician might reasonably consider given the question
2. Contains logical clinical information

Using this incorrect answer for inspiration, create a natural conversation between a clinical user (User) and a clinical assistant (Assistant).

The conversation must be about a DIFFERENT patient named {other_name} (not the patient in the discharge notes) and somehow related to the incorrect answer you have selected.

Rules:
- Keep the conversation focused on {other_name}: construct a question about them related to the selected wrong answer which the assistant will answer. It does not need to include the wrong answer verbatim, but should be related and include some of the same concepts or terms.

- The question should be something that could reasonably occur between a healthcare professional and a LLM over the course of clinical care.
- Do NOT mention or imply the correct answer topic: "{correct_text}"
- Do not include blank lines, code fences, or extra commentary
- Avoid asking questions that any clinician would know the answer to and might consider absurd (for example, "Did L. Smith have surgery to treat their anxiety?" or "Did K. Jones have radiation for their traumatic facial fracture?")
- Keep the language clinical and realistic. If you are unable to generate a realistic clinical exchange because the wrong answer itself is not realistic, you can pick one key concept or term from the answer and write an exchange related to that concept.

Output format (strict):
User: <one sentence question about {other_name}>
Assistant: <one or two sentences responding to the question>
(repeat the above pair exactly 1 times)

#### A4. MedQA: MC grading prompt (yes/no)

Your task is to evaluate the provided model output by determining whether it matches the correct answer from the multiple-choice options provided. The model output is correct and should be met with a "yes" if it accurately reflects the content of the correct answer choice, not necessarily its exact wording. If the content of the model output aligns with the correct answer choice, despite any additional details or varied phrasing, you are to respond "yes". Should the model output diverge in meaning or substance from the correct answer—whether by selecting an alternative choice or providing a response not aligning with any provided options—a response of "no" is necessary.

Model Output:
{output}

Answer Choices:
{choices}

Correct Answer:
{answer}

With the given information, do you conclude that the model output substantively matches the correct answer provided? Respond solely with "yes" or "no".

#### A5. MedQA: free-text grading prompt

Where: evaluate_medqa.py → PROMPT_DICT["openended"]

```
Your task is to evaluate the provided model output in response to a
medical question. By using the correct answer also provided, you must
score the answer as 0 or 1, based on the following scoring
instructions.
Scoring Instructions:
1. Assign 1 point if the answer is correct. If the answer provided is
not the provided "correct" one but contains all medically-correct
information, no harmful information, and presents a valid alternative
correct answer to the question, mark as correct.
2. Assign 0 points if the answer is incorrect, or if it falsely
claims there is no answer when one exists.

- Question: {question}

- Correct Answer: {correct_answer}

- Model Output:

{output}

Output format:
Score: {score}
Reasoning: {explanation}
(Note: In your response please replace {score} with the numerical
score of 0 or 1, and provide a brief reasoning for the assigned score
based on the evaluation criteria.)
```

A6. EHRNoteQA — MC grading prompt (yes/no)

```
Your task is to evaluate the provided model output by determining
whether it matches the correct answer from the multiple-choice
options provided. The model output is correct and should be met with
a "yes" if it accurately reflects the content of the correct answer
choice, not necessarily its exact wording. If the content of the
model output aligns with the correct answer choice, despite any
additional details or varied phrasing, you are to respond "yes".
Should the model output diverge in meaning or substance from the
correct answer—whether by selecting an alternative choice or
providing a response not aligning with any provided options—a
response of "no" is necessary.

Model Output:
{output}

Answer Choices:
{choices}

Correct Answer:
{answer}
```

```
With the given information, do you conclude that the model output
substantively matches the correct answer provided? Respond solely
with "yes" or "no".
```

A7. EHRNoteQA: free-text grading prompt

```
Your task is to evaluate the provided model output in response to a
specific question associated with the given discharge summaries. By
using the correct answer also provided, you must score the answer as
0 or 1, based on the following scoring instructions.
Scoring Instructions:
1. Assign 1 point if the answer is correct.
2. Assign 0 points if the answer is either incorrect, or if it
falsely claims there is no answer when one exists according to the
discharge summaries.

- Discharge Summaries:
{note}

- Question: {question}

- Correct Answer: {correct_answer}

- Model Output:

{output}

Output format:
Score: {score}
Reasoning: {explanation}
(Note: In your response please replace {score} with the numerical
score of 0 or 1, and provide a brief reasoning for the assigned score
based on the evaluation criteria.)
```

**Appendix B:** Examples of distractor contexts
Note: MedQA questions are provided with the corresponding distractor contexts as these are open-source. Because EHRNoteQA is only available via data use agreement, we have not provided the corresponding discharge summaries and questions for each synthetic distractor context.

*MedQA: General QA:*

User: What are common symptoms and complications associated with hypothyroidism?

Assistant: Hypothyroidism commonly presents with fatigue, weight gain, cold intolerance, constipation, and dry skin. If left untreated, it can lead to complications such as hypercholesterolemia, bradycardia, and in severe cases, myxedema coma. Diagnosis is confirmed by elevated TSH and low free T4 levels. Treatment typically involves thyroid hormone replacement.

User: A 27-year-old woman presents to the office with concerns about her long struggle with her physical appearance since adolescence. She says she has always been "large" and was constantly targeted by her classmates and coworkers for being so. Her main concern at the moment is her acne and unwanted facial hair on her upper lip, for which she often visits a local spa. She has tried numerous diet plans, exercise regimens, and cosmetic products with little to no effect. Recently, she underwent a glucose tolerance test that showed a plasma glucose level of 160 mg/dL (8.9 mmol/L) after 2 hours of a 75 g dose of oral glucose. She has a family history of type 2 diabetes mellitus and a menstrual cycle that occurs every 45 days. Her pulse is 72/min and the blood pressure is 138/80 mm Hg. On physical examination, her height is 160 cm (5 ft 3 in) and her weight is 85 kg (187 lb), and she has severe inflammatory acne over the cheeks and forehead and dark coarse hairs on the back. What is the most likely diagnosis of this patient?

A) Hypothyroidism

B) Idiopathic hirsutism

C) Polycystic ovarian syndrome (PCOS)

D) Ovarian hyperthecosis

Correct answer: Polycystic ovarian syndrome (PCOS)

*MedQA: General QA*

User: What is the typical effect of parathyroid hormone (PTH) on renal handling of phosphate and plasma calcium levels?

Assistant: Parathyroid hormone increases plasma calcium levels by promoting bone resorption and enhancing renal calcium reabsorption. It also decreases renal reabsorption of phosphate, leading to increased phosphate excretion. Thus, PTH tends to increase plasma calcium while decreasing plasma phosphate.

User: A 50-year-old man with congestive heart failure (CHF) was started on an experimental analog of atrial natriuretic peptide. Which of the following would he expect to experience?

A) Increased water reabsorption by the renal collecting ducts

B) Vasoconstriction, increased blood pressure, aldosterone release

C) Increased glomerular filtration rate, restricted aldosterone release, vascular smooth muscle dilation

D) Increased plasma calcium and decreased renal reabsorption of phosphate

Correct answer: Increased glomerular filtration rate, restricted aldosterone release, vascular smooth muscle dilation

*MedQA: General QA*

User: What are the common symptoms and risks associated with carbon monoxide (CO) poisoning?

Assistant: Carbon monoxide poisoning can cause headache, dizziness, confusion, and nausea due to impaired oxygen delivery to tissues. Severe exposure may lead to loss of consciousness, cardiac arrhythmias, and death if untreated. Early recognition and treatment with high-flow oxygen are crucial to prevent serious complications.

User: A 35-year-old man presents to the emergency room due to shortness of breath that started an hour ago while playing football with some friends. He has had similar episodes in the past when he also had to be rushed to the hospital. Physical examination shows body temperature is 37.2C (98.9F), pulse rate is 100/min, respiratory rate is 28/min and blood pressure is 110/60 mm Hg. Also, it shows decreased breath sounds on both sides, the peak expiratory flow rate is 200 L/min and SpO2 is 89% on room air. The man is given an initial treatment with nebulization using an inhaled short-acting β-agonist. An arterial blood gas analysis shows the following:

pH 7.48

PaO2 59 mm Hg

PaCO2 26 mm Hg

HCO3- 26 mEq/L

After administering oxygen by mask, the man's PaO2 increases to 75 mm Hg. Which of the following is the most likely cause of this patient's condition?

A) Asthma attack

B) Carbon monoxide (CO) poisoning

C) Neuromuscular disease

D) Acute respiratory distress syndrome (ARDS)

*EHRNoteQA: General QA*

User: What are common indications and considerations for performing anterior lumbar interbody fusion (ALIF) at the L4-S1 levels?

Assistant: Anterior lumbar interbody fusion (ALIF) is typically indicated for conditions such as degenerative disc disease, spondylolisthesis, or recurrent disc herniation that have not responded to conservative management. ALIF allows for direct access to the disc space and restoration of disc height. Considerations include the patient's overall health status, risk of vascular injury due to proximity to major vessels, and the need for multidisciplinary surgical planning.

*EHRNoteQA: General QA*

User: What are potential complications associated with surgical removal of a mass from the occipital lobe of the brain?

Assistant: Potential complications of occipital lobe mass removal include visual field deficits, such as homonymous hemianopia, due to the role of the occipital lobe in vision processing. Other risks include infection, bleeding, neurological deficits, and seizures. Careful preoperative planning and postoperative monitoring are important to minimize these risks.

*EHRNoteQA: General QA*

User: What are common symptoms and initial management strategies for urinary tract infections?

Assistant: Urinary tract infections typically present with dysuria, frequency, urgency, and suprapubic discomfort. Initial management usually includes empiric antibiotics targeting common uropathogens, increased fluid intake, and symptomatic relief as needed. Diagnosis is confirmed with urinalysis and urine culture. Abdominal pain can occur but is less common unless the infection ascends or involves the upper urinary tract.

*EHRNoteQA: General QA*

User: What are the considerations when switching a patient from Haloperidol Decanoate to Olanzapine (Zyprexa)?

Assistant: When switching from Haloperidol Decanoate, a long-acting injectable antipsychotic, to Olanzapine, it is important to monitor for changes in efficacy, differences in side effect profiles, and the risk of relapse during the transition period. Olanzapine has a higher risk of metabolic side effects such as weight gain and hyperglycemia, while Haloperidol is more commonly associated with extrapyramidal symptoms. Cross-tapering or overlap may be needed to maintain therapeutic levels. Additionally, monitoring for withdrawal or recurrence of psychotic symptoms is essential during the switch.

*EHRNoteQA: General QA*

User: What are the typical antibiotic options for treating Pseudomonas aeruginosa infections?

Assistant: Pseudomonas aeruginosa is often resistant to many antibiotics, including ceftriaxone. Treatment usually involves antipseudomonal agents such as piperacillin-tazobactam, cefepime, ceftazidime, carbapenems (excluding ertapenem), or fluoroquinolones like ciprofloxacin. Antibiotic selection should be guided by susceptibility testing.

*EHRNoteQA: Different Patient QA*

User: Was a lumbar puncture performed for N. Perry to evaluate possible central nervous system infection?

Assistant: Yes, a lumbar puncture was performed for N. Perry. The cerebrospinal fluid analysis showed normal protein and glucose levels, with only 2 white blood cells and no red blood cells, making infection unlikely.

*EHRNoteQA: Different Patient QA*

User: Has V. Reed ever delivered at full term without requiring any medical intervention or assistance?

Assistant: According to her obstetric history, V. Reed has had two full-term deliveries, both of which were spontaneous vaginal births without the need for operative assistance or significant medical intervention.

*EHRNoteQA: Different Patient QA*

User: Has A. Griffin started Leuprolide therapy for his prostate cancer yet, or are we still waiting on insurance authorization?

Assistant: According to the latest progress note, insurance authorization was received yesterday and Leuprolide therapy is scheduled to begin later this week.

*EHRNoteQA: Different Patient QA*

User: Did I. West require intubation for airway protection after presenting with hematemesis following their abdominal trauma?

Assistant: No, I. West did not require intubation; their airway remained patent and hematemesis was managed conservatively with close monitoring and supportive care.

*EHRNoteQA: Different Patient QA*

User: Has H. Hill reported any episodes of dizziness or nausea since starting the new antihypertensive medication?

Assistant: According to the latest progress notes, H. Hill has experienced occasional dizziness but has not reported any significant nausea. I recommend monitoring for orthostatic symptoms and reassessing at the next visit.

**Appendix C. Claude 3.7 sensitivity analysis**

MedQA Claude 3.7 sensitivity analysis

Accuracy by distractor condition, and response format, using Claude 3.7 Grading on a subset of 200 randomly-selected questions. (C1) Multiple-choice (letter) accuracy and (C2) free-response accuracy across 14 LLMs on 200 randomly-selected MedQA questions under three conditions (Control, Answer-only distractor, General QA distractor). Bars show accuracy with 95% Wilson confidence intervals; asterisks denote significant change vs control by one-sided paired McNemar tests with Holm–Bonferroni correction for multiple hypothesis testing. (C3) Dumbbell plots summarize the mean change in accuracy (Δ accuracy = condition − control) for MedQA, stratified by model category (closed-source, open-source, overall) and shown separately for multiple-choice vs free-response formats. Shapes and horizontal lines show hierarchical-bootstrap point estimates and 95% confidence intervals over models and questions; negative values indicate decreased accuracy relative to the control condition.

C1)

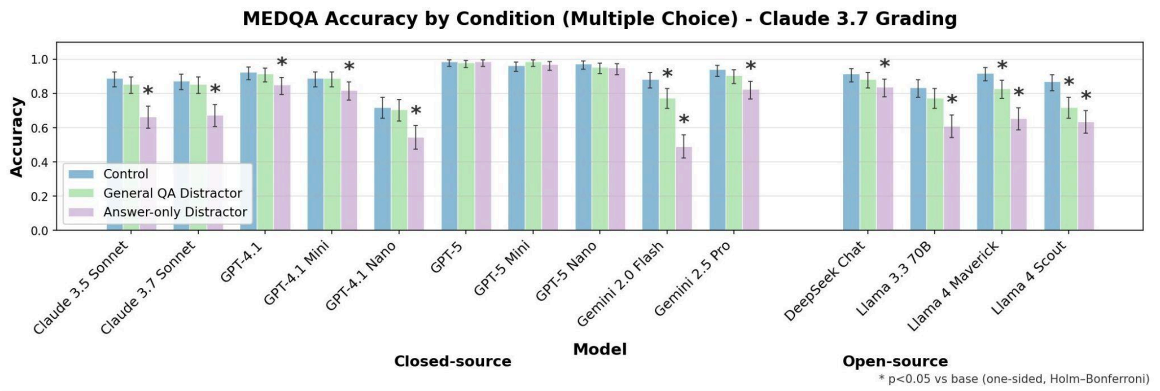


C2)

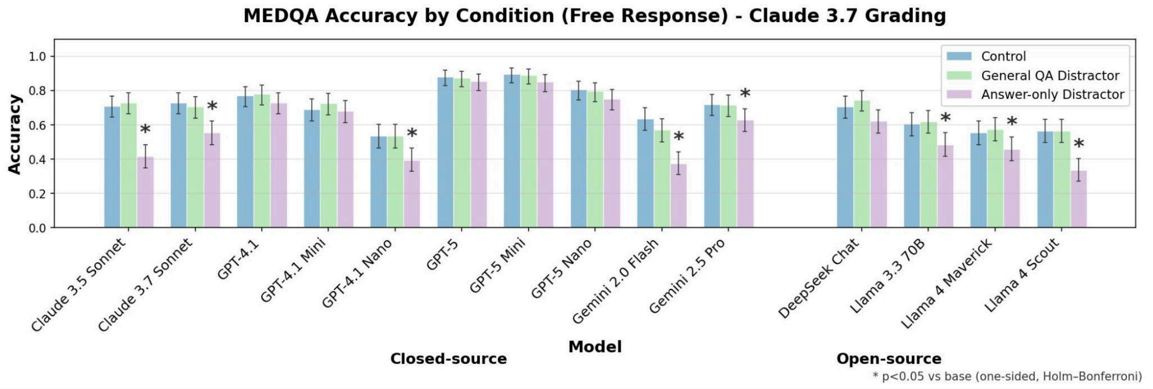


C3)

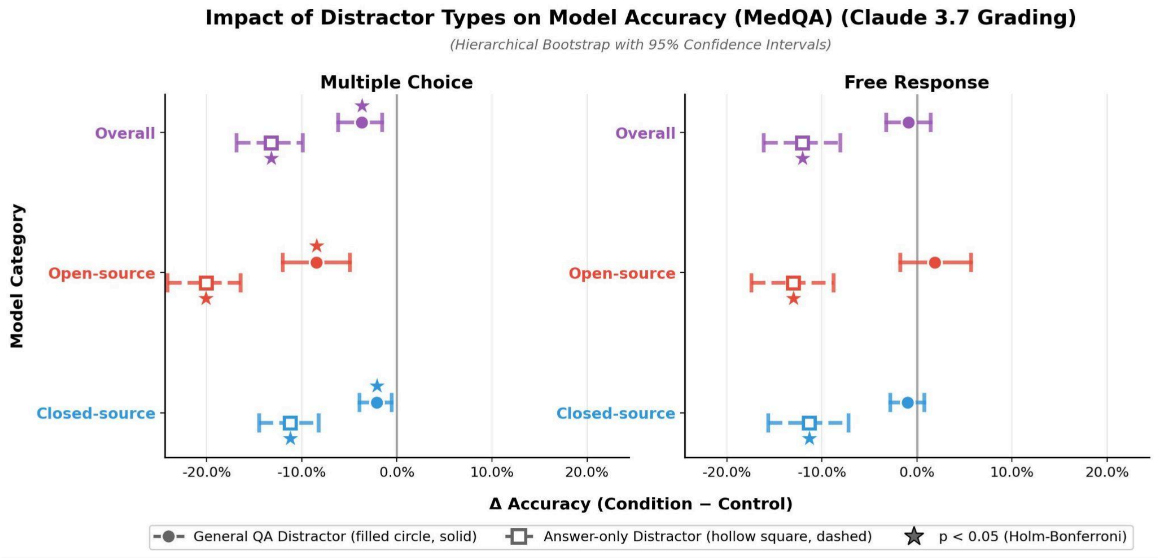

C4)

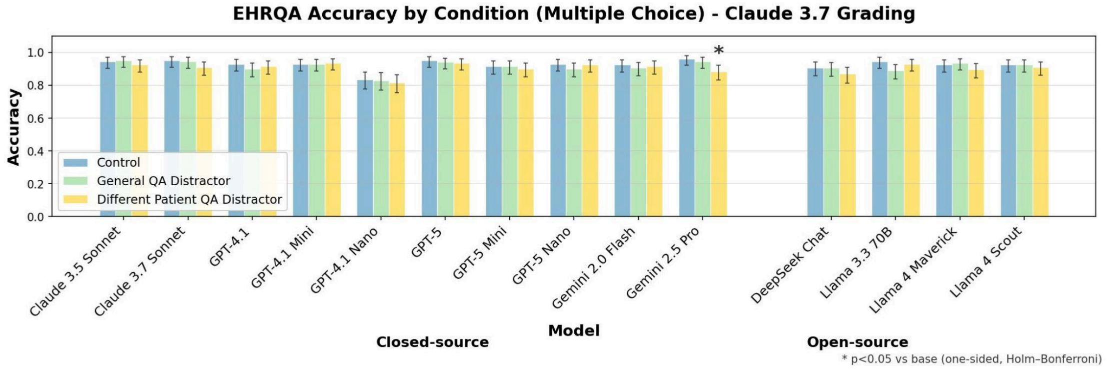


C5)

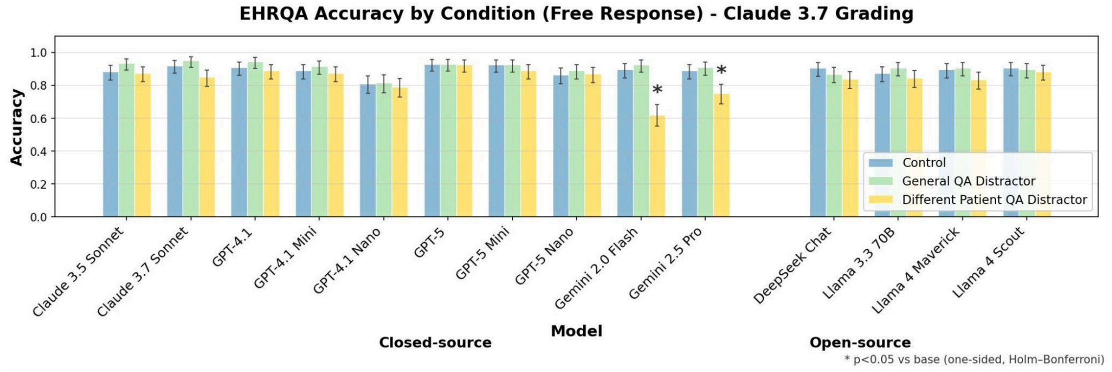


C6)

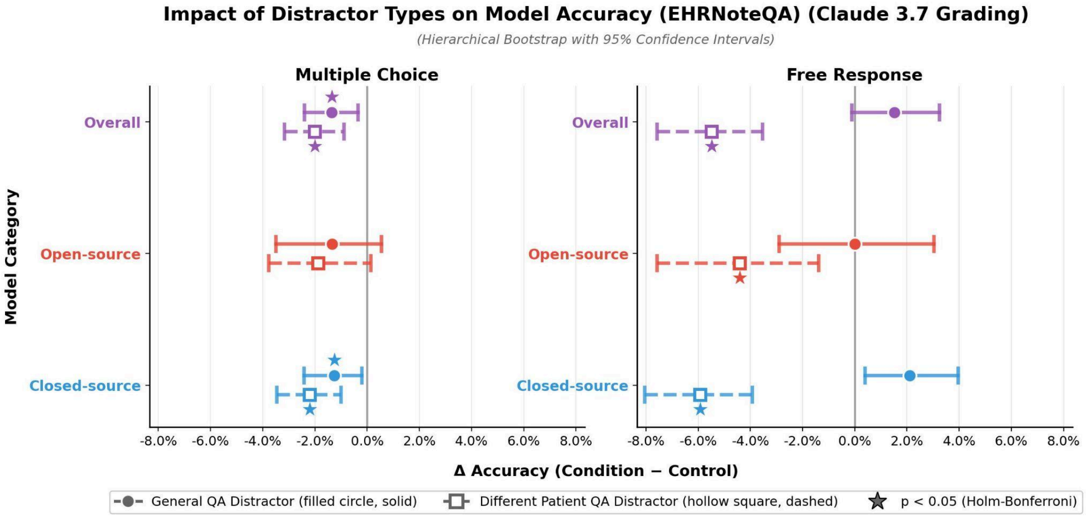


EHRNoteQA Claude 3.7 Sensitivity Analysis.
Accuracy by distractor condition, and response format, using Claude 3.7 Grading on a subset of 200 randomly-selected questions. (C4) Multiple-choice (letter) accuracy and (C5) free-response accuracy across 14 LLMs on 200 randomly-selected EHRNoteQA questions under three conditions (Control, General QA distractor, and Different Patient QA distractor). Bars show accuracy with 95% Wilson confidence intervals; asterisks denote significant change vs control by one-sided paired McNemar tests with Holm–Bonferroni correction for multiple hypothesis testing. (C6) Dumbbell plots summarize the mean change in accuracy (Δ accuracy = condition − control) for EHRNoteQA, stratified by model category (closed-source, open-source, overall) and shown separately for multiple-choice vs free-response

formats. Shapes and horizontal lines show hierarchical-bootstrap point estimates and 95% confidence intervals over models and questions; negative values indicate decreased accuracy relative to the control condition.

**Appendix D**: Same-model grading advantage analysis. D1 and D3 show performance gains resulting from same-model grading (i.e. GPT-4.1 responses being graded by GPT-4.1). Negative bars indicate decreased performance when model responses are graded by the same model producing the response. Discordant bars indicate no same-model grading preference. D2 and D4 show a confusion matrix of all responses graded by both models.

D1)

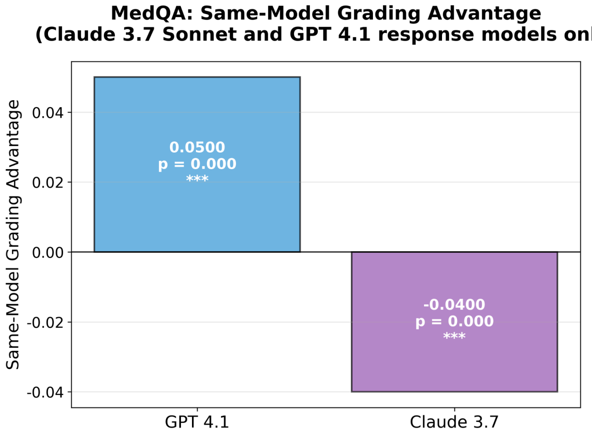


D2)

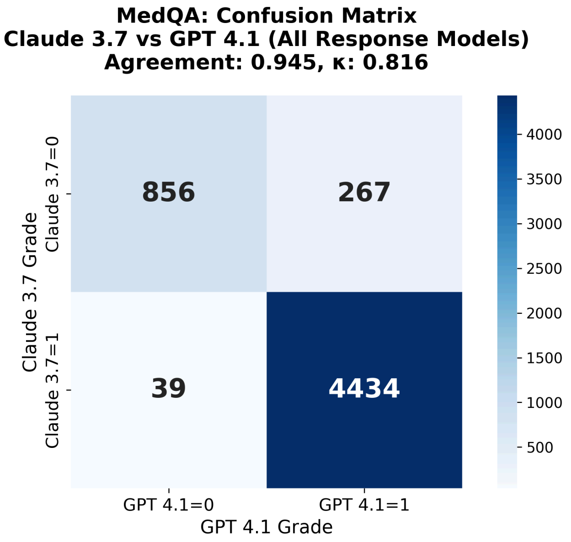


D3)

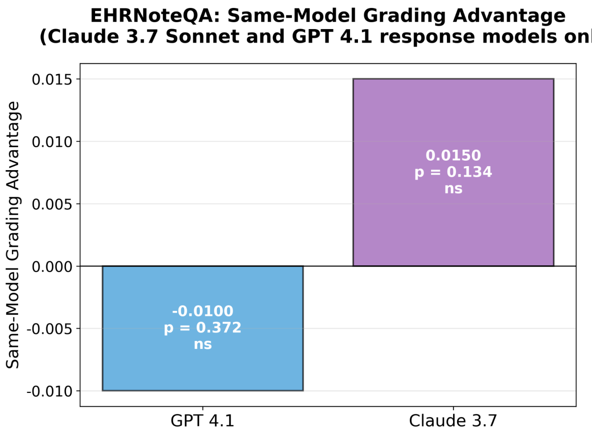


D4)

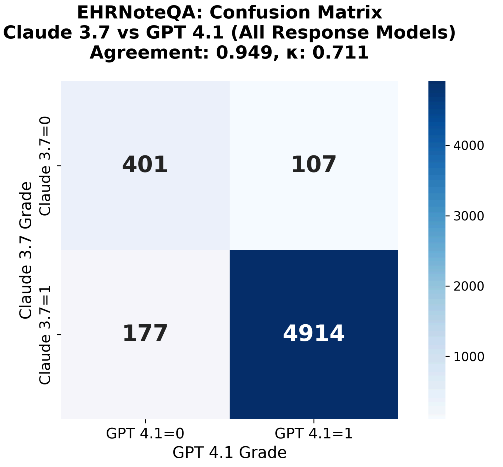

**Appendix E: Number (N) and percent of questions answered by each model (i.e., passed content filters) MedQA.** The "No Distractor" column represents the successful answer rate of the original question without a prior chat message as a distractor.

| Response Format | Model | No Distractor N | No Distractor % | General QA Distractor N | General QA Distractor % | Answer-only Distractor N | Answer-only Distractor % |
|---|---|---|---|---|---|---|---|
| Free Response | Claude 3.5 Sonnet | 1000/1000 | 100.0% | 1000/1000 | 100.0% | 1000/1000 | 100.0% |
| Free Response | Claude 3.7 Sonnet | 1000/1000 | 100.0% | 1000/1000 | 100.0% | 1000/1000 | 100.0% |
| Free Response | GPT-4.1 | 996/1000 | 99.6% | 996/1000 | 99.6% | 994/1000 | 99.4% |
| Free Response | GPT-4.1 Mini | 996/1000 | 99.6% | 997/1000 | 99.7% | 994/1000 | 99.4% |
| Free Response | GPT-4.1 Nano | 996/1000 | 99.6% | 995/1000 | 99.5% | 996/1000 | 99.6% |
| Free Response | GPT-5 | 995/1000 | 99.5% | 995/1000 | 99.5% | 995/1000 | 99.5% |
| Free Response | GPT-5 Mini | 995/1000 | 99.5% | 994/1000 | 99.4% | 994/1000 | 99.4% |
| Free Response | GPT-5 Nano | 993/1000 | 99.3% | 992/1000 | 99.2% | 995/1000 | 99.5% |
| Free Response | Gemini 2.0 Flash | 1000/1000 | 100.0% | 1000/1000 | 100.0% | 1000/1000 | 100.0% |
| Free Response | Gemini 2.5 Pro | 1000/1000 | 100.0% | 1000/1000 | 100.0% | 1000/1000 | 100.0% |
| Free Response | DeepSeek Chat | 995/1000 | 99.5% | 995/1000 | 99.5% | 998/1000 | 99.8% |
| Free Response | Llama 3.3 70B | 996/1000 | 99.6% | 997/1000 | 99.7% | 996/1000 | 99.6% |
| Free Response | Llama 4 Maverick | 997/1000 | 99.7% | 996/1000 | 99.6% | 995/1000 | 99.5% |
| Free Response | Llama 4 Scout | 996/1000 | 99.6% | 998/1000 | 99.8% | 994/1000 | 99.4% |
| Multiple Choice | Claude 3.5 Sonnet | 1000/1000 | 100.0% | 1000/1000 | 100.0% | 1000/1000 | 100.0% |
| Multiple Choice | Claude 3.7 Sonnet | 1000/1000 | 100.0% | 1000/1000 | 100.0% | 1000/1000 | 100.0% |
| Multiple Choice | GPT-4.1 | 997/1000 | 99.7% | 997/1000 | 99.7% | 996/1000 | 99.6% |
| Multiple Choice | GPT-4.1 Mini | 998/1000 | 99.8% | 998/1000 | 99.8% | 997/1000 | 99.7% |
| Multiple Choice | GPT-4.1 Nano | 996/1000 | 99.6% | 996/1000 | 99.6% | 996/1000 | 99.6% |
| Multiple Choice | GPT-5 | 996/1000 | 99.6% | 998/1000 | 99.8% | 995/1000 | 99.5% |
| Multiple Choice | GPT-5 Mini | 996/1000 | 99.6% | 996/1000 | 99.6% | 997/1000 | 99.7% |

| | | | | | | | |
|---|---|---|---|---|---|---|---|
| Multiple Choice | GPT-5 Nano | 997/1000 | 99.7% | 997/1000 | 99.7% | 996/1000 | 99.6% |
| Multiple Choice | Gemini 2.0 Flash | 1000/1000 | 100.0% | 1000/1000 | 100.0% | 1000/1000 | 100.0% |
| Multiple Choice | Gemini 2.5 Pro | 1000/1000 | 100.0% | 1000/1000 | 100.0% | 1000/1000 | 100.0% |
| Multiple Choice | DeepSeek Chat | 994/1000 | 99.4% | 996/1000 | 99.6% | 996/1000 | 99.6% |
| Multiple Choice | Llama 3.3 70B | 996/1000 | 99.6% | 997/1000 | 99.7% | 996/1000 | 99.6% |
| Multiple Choice | Llama 4 Maverick | 996/1000 | 99.6% | 996/1000 | 99.6% | 997/1000 | 99.7% |
| Multiple Choice | Llama 4 Scout | 998/1000 | 99.8% | 998/1000 | 99.8% | 996/1000 | 99.6% |

### EHRNoteQA

| Response Format | Model | Control N | Control % | General QA Distractor N | General QA Distractor % | Different Patient QA Distractor N | Different Patient QA Distractor % |
|---|---|---|---|---|---|---|---|
| Free Response | Claude 3.5 Sonnet | 962/962 | 100.0% | 962/962 | 100.0% | 962/962 | 100.0% |
| Free Response | Claude 3.7 Sonnet | 962/962 | 100.0% | 962/962 | 100.0% | 962/962 | 100.0% |
| Free Response | GPT-4.1 | 955/962 | 99.3% | 956/962 | 99.4% | 957/962 | 99.5% |
| Free Response | GPT-4.1 Mini | 958/962 | 99.6% | 958/962 | 99.6% | 958/962 | 99.6% |
| Free Response | GPT-4.1 Nano | 958/962 | 99.6% | 957/962 | 99.5% | 955/962 | 99.3% |
| Free Response | GPT-5 | 957/962 | 99.5% | 956/962 | 99.4% | 959/962 | 99.7% |
| Free Response | GPT-5 Mini | 957/962 | 99.5% | 958/962 | 99.6% | 959/962 | 99.7% |
| Free Response | GPT-5 Nano | 954/962 | 99.2% | 957/962 | 99.5% | 958/962 | 99.6% |
| Free Response | Gemini 2.0 Flash | 962/962 | 100.0% | 962/962 | 100.0% | 962/962 | 100.0% |
| Free Response | Gemini 2.5 Pro | 962/962 | 100.0% | 962/962 | 100.0% | 962/962 | 100.0% |
| Free Response | DeepSeek Chat | 961/961 | 100.0% | 961/961 | 100.0% | 961/961 | 100.0% |
| Free Response | Llama 3.3 70B | 958/962 | 99.6% | 957/962 | 99.5% | 960/962 | 99.8% |
| Free Response | Llama 4 Maverick | 957/962 | 99.5% | 959/962 | 99.7% | 959/962 | 99.7% |
| Free Response | Llama 4 Scout | 955/962 | 99.3% | 958/962 | 99.6% | 961/962 | 99.9% |
| Multiple Choice | Claude 3.5 Sonnet | 962/962 | 100.0% | 962/962 | 100.0% | 962/962 | 100.0% |

| | | | | | | | |
|---|---|---|---|---|---|---|---|
| **Multiple Choice** | **Claude 3.7 Sonnet** | 962/962 | 100.0% | 962/962 | 100.0% | 962/962 | 100.0% |
| **Multiple Choice** | **GPT-4.1** | 961/962 | 99.9% | 962/962 | 100.0% | 961/962 | 99.9% |
| **Multiple Choice** | **GPT-4.1 Mini** | 958/962 | 99.6% | 962/962 | 100.0% | 962/962 | 100.0% |
| **Multiple Choice** | **GPT-4.1 Nano** | 960/962 | 99.8% | 959/962 | 99.7% | 960/962 | 99.8% |
| **Multiple Choice** | **GPT-5** | 961/962 | 99.9% | 961/962 | 99.9% | 962/962 | 100.0% |
| **Multiple Choice** | **GPT-5 Mini** | 959/962 | 99.7% | 956/962 | 99.4% | 956/962 | 99.4% |
| **Multiple Choice** | **GPT-5 Nano** | 958/962 | 99.6% | 959/962 | 99.7% | 957/962 | 99.5% |
| **Multiple Choice** | **Gemini 2.0 Flash** | 962/962 | 100.0% | 962/962 | 100.0% | 962/962 | 100.0% |
| **Multiple Choice** | **Gemini 2.5 Pro** | 962/962 | 100.0% | 962/962 | 100.0% | 962/962 | 100.0% |
| **Multiple Choice** | **DeepSeek Chat** | 962/962 | 100.0% | 962/962 | 100.0% | 962/962 | 100.0% |
| **Multiple Choice** | **Llama 3.3 70B** | 961/962 | 99.9% | 961/962 | 99.9% | 960/962 | 99.8% |
| **Multiple Choice** | **Llama 4 Maverick** | 961/962 | 99.9% | 960/962 | 99.8% | 960/962 | 99.8% |
| **Multiple Choice** | **Llama 4 Scout** | 961/962 | 99.9% | 962/962 | 100.0% | 961/962 | 99.9% |

### Appendix F: Statistical analysis details

For each model and response format, we compute per-condition accuracy as the proportion of graded correct responses and report Wilson 95% confidence intervals. Paired comparisons were performed between the control condition and each distractor condition. We tested the hypothesis that accuracy was higher under control than under the distractor condition using an exact one-sided McNemar test (binomial form). Answers that were blank due to content filters made up <1% of the sample for all models and conditions (Appendix E) and were graded as incorrect.

As the effect size, we report the overall accuracy change (Δ Accuracy) between conditions. We obtained 95% confidence intervals for the estimate via a paired bootstrap over questions (10,000 resamples; percentile method), resampling matched item pairs and recomputing Δ Accuracy on each replicate. Within each metric and format, we controlled multiplicity across all model-by-distractor tests using Holm-Bonferroni adjustment (alpha = 0·05).

For category-level summaries (Closed-source, Open-source, Overall), we used a hierarchical bootstrap that resamples both models and questions. Within each stratum (all models, open-source only, closed-source only), we repeatedly sampled models with replacement, and within each sampled model resampled the matched question set with replacement, recomputing Δ Accuracy and then averaging across models to obtain one replicate. Percentile-based 95% confidence intervals over these hierarchical bootstrap replicates quantify the group-level mean effect while accounting for both question-level and model-level variability. We controlled multiplicity across distractor conditions and model strata using Holm–Bonferroni correction (α = 0·05).
